# Childhood trauma is prevalent and associated with co-occurring depression, anxiety, mania and psychosis in young people attending Australian youth mental health services

**DOI:** 10.1101/2023.03.16.23287385

**Authors:** Sarah Bendall, Oliver Eastwood, Tim Spelman, Patrick McGorry, Ian Hickie, Alison R. Yung, Paul Amminger, Stephen J. Wood, Christos Pantelis, Rosemary Purcell, Lisa Phillips

**Author notes:** **Corresponding author:** Sarah Bendall, Orygen, 35 Poplar Rd, Parkville, VIC 3052, Australia.

## Abstract

**Objectives:** Childhood trauma is common and associated with mental ill health. While high rates of trauma are observed across individual disorder groups, there is evidence that trauma is associated with an admixture of affective, anxiety, and psychotic symptoms in adults. Given that both early onset of mental disorder and trauma exposure herald poor outcomes, it is important to examine trauma prevalence rates in youth mental health services and to determine whether this trauma-related clustering is present in help-seeking young people.

**Methods:** We used data from the Transitions Study, a longitudinal investigation of young people attending one of four headspace youth mental health services in Melbourne and Sydney, Australia between January 2011 and August 2012. Participants were 775 young people aged 12 to 25 (65.9% female; mean age = 18.3, SD = 3.2). Childhood trauma was assessed using the Childhood Trauma Questionnaire (CTQ). Multinomial regression was used to assess whether reported childhood trauma was more strongly associated with the co-occurrence of depression, anxiety, mania, and psychosis symptoms than with any one in isolation.

**Results:** Approximately 84% of participants reported some form of abuse (emotional: 68%; physical: 32%; sexual: 22%) or neglect (emotional: 65%; physical: 46%) during childhood or adolescence. Exposure to multiple trauma types was common. Childhood trauma was significantly associated with each symptom domain (depression, mania, anxiety and psychosis). Childhood trauma was more strongly associated with the co-occurrence of these symptoms than with any one of these domains in isolation, such that trauma-exposed young people were more likely to experience increased symptom clustering than their non-exposed counterparts.

**Conclusions:** Childhood trauma is pervasive in youth mental health services and associated with a heterogeneous symptom profile that cuts across traditional diagnostic boundaries.

---

Exposure to childhood traumas, such as experiencing or witnessing physical or sexual abuse, is common and associated with a range of deleterious mental health outcomes. General population estimates suggest that more than two thirds of young people will experience at least one traumatic event by age 16 (Copeland et al., 2007); the majority of whom will experience multiple events (McLaughlin et al., 2012). Such traumas reportedly account for 45% of the population attributable risk for childhood-onset mental disorders and 32% for adolescent-onset mental disorders (Green et al., 2010). Prospective studies have implicated childhood trauma in suicide and the development of a range of psychopathologies, including affective, psychotic, substance abuse, post-traumatic stress, anxiety symptoms, and personality disorders (Spataro et al., 2004; Widom, 1999; Widom et al., 2007; Cutajar et al., 2010). These findings are underscored by meta-analytic research demonstrating robust associations between childhood trauma and psychosis (Varese et al., 2012), anxiety (Fernandes and Osorio, 2015), depression (Mandelli et al., 2015), and bipolar disorder (Palmier-Claus et al., 2016).

Contemporary youth mental health service models often cater for young people across a broad diagnostic and severity spectrum of mental health disorders (McGorry et al., 2013). If services are to respond to these challenges and provide appropriate intervention for the effects of trauma, the prevalence of trauma exposure in the young people they serve must be known. While high rates of trauma are noted in first-episode psychosis services (Duhig et al., 2015; Peach et al., 2021; Trauelsen et al., 2015), and in youth inpatient psychiatric settings (Allwood et al., 2008; Havens et al., 2012), general prevalence rates among help-seeking youth presenting for different mental health concerns of varying severity are not well known. This information is necessary to drive appropriate service delivery. For example, high rates of trauma would indicate need for trauma screening and assessment protocols and guidelines for intervention.

In addition to knowing prevalence of trauma exposure, it is also important to understand its role in symptom severity and complexity in help-seeking young people. In those with psychosis, childhood trauma is associated with worse functional outcomes (Alameda et al., 2015) and more severe hallucinations and delusions compared to non-abused counterparts (Bailey et al., 2018). Childhood maltreatment also increases the risk of development of psychotic disorder and poor functional outcomes in youth at risk for psychosis (Thompson et al., 2014; Yung et al., 2015). Maltreated individuals with depressive, substance use, and anxiety disorders have been shown to have earlier disorder onset, greater symptom severity, poorer quality of life, increased suicidality, and worse treatment response compared to disordered individuals without trauma histories (Teicher and Samson, 2013).

Recent longitudinal work suggests that individuals shift between separate disorder types (internalising, externalizing, and thought disorders) across the lifespan, rather than ‘sticking’ with the one disorder (Caspi et al., 2020). As such, childhood trauma may portend diverse mental health symptomology rather than increase risk for discrete disorders. Indeed, in a clinical sample of 162 adult outpatients, childhood abuse was shown to predict increased symptom complexity, as measured by simultaneous elevated clinical scales on the Minnesota Multiphasic Personality Inventory (Choi et al., 2014). In a representative sample of adults, Van Nierop and colleagues (2015) found childhood trauma to be more strongly associated with an increased lifetime clustering of symptoms—affective, psychotic, anxiety-related, and manic—than with any one of these domains in isolation. In other words, those with trauma histories were more likely to experience multiple mental health symptoms across the lifespan compared to their non-abused counterparts. This pattern of results was observed in three clinical samples (adults with a lifetime mood, anxiety, or non-affective psychotic disorder), and a sub-sample of non-help-seeking, non-diagnosed individuals (van Nierop et al., 2015).

Given that both trauma exposure and early onset of disorder compound mental health outcomes (Caspi et al., 2020; Teicher and Samson, 2013), it is important to examine the relationship between trauma and symptom clustering in help-seeking young people. Trauma-related symptom clustering in youth would provide evidence that this symptom profile is already present in earlier expressions of psychopathology and indicate the need for earlier intensive and trans-diagnostic care. Indeed, in a follow-up study, Van Nierop and colleagues (2016) showed trauma-related symptom co-occurrence to have functional and clinical relevance (reduced quality of life, greater incidence of substance use disorders, lower global functioning, and more help-seeking behaviours).

The present study had two aims. First, to examine the prevalence of childhood trauma among help-seeking young people accessing an Australian youth mental health care service. Second, to examine associations between childhood trauma and mental health symptoms, and to determine whether a previously reported association between childhood trauma and affective, anxiety, and psychosis symptom admixture in adults is present in a help-seeking youth sample.

## Method

The present study employs data from the Transitions Study, a prospective investigation of help-seeking young people attending one of four *headspace* youth mental health services in Melbourne and Sydney, Australia between January 2011 and August 2012. The study methodology has been described in detail elsewhere (Purcell et al., 2015a).

### Participants

As headspace centres focus both on youth mental health and early intervention, young people may present for care with varying illness severity, with an emphasis on mild to moderate severity, across a range of mental health problems. Participants from the Transitions Study were included here if they completed the trauma assessment instrument. Of the total cohort of 801, 775 young people completed the trauma assessment and were included in the current study. Participants were aged 12 to 25 (65.9% female; mean age = 18.3, SD = 3.2). For further description of demographic details of the full Transitions sample, see Purcell and colleagues (2015b). All participants were English-speaking and able to provide informed consent when approached for recruitment. There were no exclusion criteria other than significant intellectual disability (e.g. IQ < 65) that would preclude the ability to provide informed consent and complete the study assessment.

### Measures

#### Quick Inventory of Depressive Symptomatology (QIDS) 16-item adolescent version

The QIDS (Rush et al., 2003) assesses the presence, during the previous seven days, of the major diagnostic symptoms of depression according to the Diagnostic and Statistical Manual of Mental Disorders, Fourth Edition. Symptoms are rated on a 4-point Likert scale and combined to provide total scores ranging from 0 to 27. Depression presence was defined by a total score of 7 or more.

#### Young Mania Rating Scale (YMRS)

The YMRS 11-item measure indicates the nature and severity of manic symptoms within the past 48 hours (Young et al., 1978). Each item is graded across five explicitly defined anchor points (ranging from 0–4 for seven items to 0–8 for four items). The rating of items is based both on subjective report by the participant and the interviewer’s behavioural observations. Total scores range from 0 to 60. Mania presence was defined by a total score of 20 or more.

#### Comprehensive Assessment of the At-Risk Mental State (CAARMS)

The CAARMS is a semi-structured interview that assesses the presence and severity of psychotic symptoms over the past 12 months (Yung et al., 2005). The Positive Symptom Scale was used in this study and consists of four subscales: (i) unusual thought content; (ii) non-bizarre ideas; (iii) perceptual abnormalities; and (iv) disorganised speech. Psychosis presence was defined by: a global rating score of 6 on unusual thought content, non-bizarre ideas or disorganized speech; or a rating of 5–6 on perceptual abnormalities; *and* an associated frequency score of 4–6, *and* with symptoms lasting 1 week or longer.

#### Generalized Anxiety Disorder scale 7-item version (GAD-7)

The GAD-7 measures core symptoms of generalized anxiety disorder (e.g. feeling nervous, unable to relax, worrying about different things, afraid something awful might happen (Spitzer et al., 2006). Participants rated the frequency with which they have experienced these anxiety symptoms in the past 2 weeks on a 4-point Likert scale. Anxiety presence was defined by a total score of 10 or more.

#### Childhood Trauma Questionnaire (CTQ)

The presence of childhood trauma was measured by the CTQ (Bernstein et al., 1994). The CTQ is a self-report 28-item retrospective scale that measures the occurrence of abuse (emotional, physical, and sexual) and neglect (emotional and physical) in childhood and adolescence. The CTQ is a widely used measure, and its psychometric properties have been extensively assessed (Bernstein et al., 1997). A 3-item scale is also used to detect false-negative trauma reports (e.g. ‘I had the perfect childhood’). Participants indicated the extent to which they experienced each item while they were ‘growing up’ according to a 5-point Likert scale. Childhood emotional; physical, sexual abuse; emotional and physical neglect was each classified as present or absent on the basis of widely used cut scores with high sensitivity and acceptable specificity for the prevalence analysis (Bernstein et al., 1997). An overall dichotomous total childhood trauma factor was derived: the no trauma group included all those who had been classified as having the absence of each of the five childhood trauma sub-scales; the trauma group consisted of those who had been classified as having trauma present in at least one of the five trauma sub-scales. Trauma presence was further categorised according to severity (low, moderate, and severe) across each trauma sub-scale (Bernstein et al., 1997). Total CTQ scores were modeled as continuous variables when exploring symptomatic correlates of childhood trauma and its relationship with isolated *v*. multiple symptom domains. Total CTQ scores rather than sub-scales were used as sub-scales are known to be intercorrelated and empirical and theoretical evidence suggests that trauma persistence and severity rather than specific trauma exposures confer risk for mental disorder (Teicher, 2012; Trauelsen et al., 2015).

### Procedure

The study protocol was approved by Human Research Ethics Committees at the University of Melbourne and the University of Sydney. Participants aged 15 years and over provided written informed consent, whereas those aged 12–14 years (inclusive) assented with written informed consent provided by a parent or guardian. Research assistants (RA) with a minimum 4-year graduate psychology degree implemented the study protocol. The RAs were trained in the use of each study measure and achieved an interrater reliability score of at least 0.8 on each of the interviewer-rated clinical measures before recruitment commenced. The RAs conducted structured interviews with each participant using the clinical measures described above (QIDS, YMRS, CAARMS, GAD-7) before then providing an iPad or laptop for the completion of a self-report questionnaire (CTQ) and demographics. Assessment took approximately 1.5 to 2 hours to complete and participants were compensated with a $20 gift voucher for their time.

### Statistical analyses

Categorical variables were summarized using frequency and percentage. Continuous variables were summarized using mean and standard deviation (SD) or median and inter-quartile range (IQR) as appropriate. Spearman rank correlations were used to quantify associations between childhood trauma and isolated symptom domains (mania, depression, anxiety). CTQ scores were compared between the psychosis and non-psychosis groups using a Wilcoxon rank-sum test. We followed the statistical approach outlined by Van Nierop et al. (2015) to investigate whether childhood trauma was associated with isolated or co-occurring mental health symptoms in help-seeking young people. Multinomial logit regression analyses were conducted and effect sizes were obtained with post-hoc linear combination of estimators (lincom) tests and expressed as ratios of Odds Ratios, adjusting for age and sex. The multinomial outcome variable was comprised of the following categories: (1) no symptoms, (2) depression only (based on QIDS score as defined above), (3) anxiety only (based on GAD-7 score), (4) positive psychotic symptoms only (based on CAARMS score), (5) a combination of two symptoms domains (any combination of depression, mania, anxiety or psychosis), (6) a combination of three symptom domains, (7) all four symptom domains combined. These outcome variables differ from those used by Van Nierop et al. (2015), in that there is no ‘mania only’ variable. Initial univariate models of total CTQ score predicting each of the four separate symptoms were run to ensure there was sufficient power for each domain under consideration. CTQ score significantly predicted all symptoms except the mania domain, which was likely secondary to the small number of young people satisfying the YRMS criteria. A mania only variable was not included for this reason. Goodness of fit for the logit models was assessed using a Hosmer & Lemeshow test. All analyses were conducted using Stata version 15 (StataCorp, 2017).

## Results

### Participant characteristics and childhood trauma prevalence

Participant demographics and characteristics are presented in Table 1. Approximately 84% (n = 645) of participants reported exposure to some form of childhood trauma (low, moderate, severe). Of the total group, 20% (n= 151) of participants scored in the moderate category for at least one of the sub-types of trauma and 39% (n= 305) scored in the severe range in at least one of the trauma sub-categories. Trauma prevalence and severity for each trauma type are presented in Figure 1. The most commonly reported type of trauma was emotional abuse (68%, n =527) and the least common sexual abuse (22%, n= 173). Poly-traumatisation was common, with 67.1% (n= 520) of participants reporting exposure to more than one form of trauma (see Table 2), 72.0% (n= 558) reporting any form of abuse and 71.1% (n= 551) reporting any form of neglect.

**Table 1.** Participant demographics and characteristics

| Variable | Value (*n* = 775) |
| --- | --- |
| Gender: n=female | 511 (65.9%) |
| Age | 18.3 (3.2) |
| Aboriginal or Torres Strait Islander | 4% |
| Depression: QIDS<sup>a</sup> total score | 10.2 (5.3) |
| Mania: YMRS<sup>b</sup> total score | 3.9 (4.8) |
| Anxiety: GAD-7<sup>c</sup> total score | 9.9 (6.0) |
| Psychosis: % meeting CAARMS criteria | 7.2% |
| Childhood trauma: CTQ total score | 47.0 (17.1) |

Missing data: a = 1; b = 2; c = 4
Values given are mean and (S.D.) unless otherwise stated. CAARMS, Comprehensive Assessment of At-Risk Mental States; CTQ, Childhood Trauma Questionnaire; GAD, Generalized Anxiety Disorder; OASIS, Overall Anxiety Severity and Impairment Scale; QIDS, Quick Inventory of Depressive Symptomatology; SD, standard deviation; YMRS, Young Mania Rating Scale

**Table 2.** Cumulative trauma exposure

| Number of trauma types (CTQ) | *n* (%) |
| --- | --- |
| 0 | 126 (16.3) |
| 1 | 129 (16.7) |
| 2 | 152 (19.6) |
| 3 | 164 (21.2) |
| 4 | 139 (17.9) |
| 5 | 65 (8.4) |

**Figure 1.**
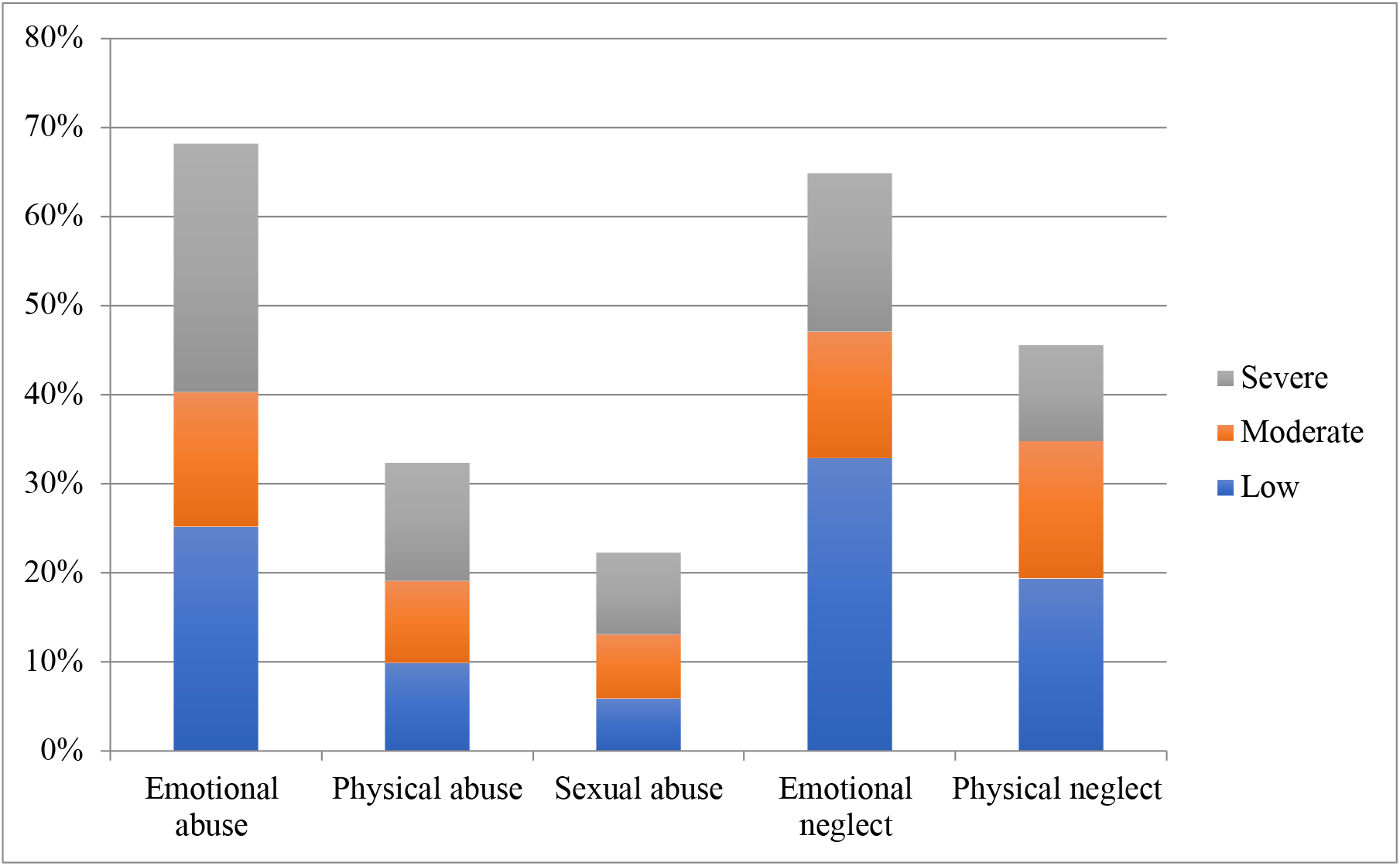
Childhood trauma prevalence and severity by trauma type.

### Associations of childhood trauma with separate symptoms

Table 3 presents associations between childhood trauma and isolated symptom domains. There was a small positive correlation between total CTQ scores and QIDS, GAD-7, YMRS scores individually, such that higher scores on the CTQ were associated with higher scores on the symptom measures. Mean CTQ scores were significantly higher in the psychosis group (M= 53.98, SD= 18.14) compared to the non-psychosis group (M= 46.46, SD= 16.88, *p* < .001).

**Table 3.** Associations between symptom scores domains and childhood trauma (CTQ total)

|  | R (Spearman's) | P |
| --- | --- | --- |
| QIDS (depression) | 0.2620 | < 0.01 |
| GAD-7 (anxiety) | 0.2289 | < 0.01 |
| YRMS (mania) | 0.1136 | 0.0016 |

### Associations of childhood trauma with multiple symptoms

Table 4 shows odds ratios regarding whether childhood trauma was associated with an increased likelihood of multiple symptom domains, rather than association with single symptom domains (e.g. association with psychosis symptoms in isolation versus association with psychosis symptoms co-occurring with any combination of 1, 2 or 3 symptoms of mania, depression or anxiety). There is a pattern of significantly greater odds ratios as more symptom domains were present. Childhood trauma was thus more strongly associated with increasing combinations of depression, anxiety, mania, and psychosis symptoms, than any one of the isolated symptom domains.

**Table 4.** Associations of childhood trauma and isolated symptoms versus clustering of symptoms

|  | v. depression<br>ROR (95% CI) | v. anxiety<br>ROR (95% CI) | v. psychosis<br>ROR (95% CI) | v. 2 symptom<br>domains ROR<br>(95% CI) | v. 3 symptom<br>domains ROR<br>(95% CI) |
| --- | --- | --- | --- | --- | --- |
| 2<br>symptom<br>domains | 1.05*** (1.03<br>– 1.08) | 1.05*** (1.02<br>– 1.08) | 1.07 (1 – 1.14) |  |  |
| 3<br>symptom<br>domains | 1.06*** (1.03<br>– 1.09) | 1.06*** (1.03<br>– 1.1) | 1.08* (1.01 –<br>1.15) | 1.08*** (1.05 –<br>1.11) |  |
| 4<br>symptom<br>domains | 1.11*** (1.05<br>– 1.16) | 1.10*** (1.05<br>– 1.16) | 1.12** (1.04 –<br>1.22) | 1.12*** (1.07 –<br>1.18) | 1.13*** (1.07<br>– 1.19) |

ROR = ratio of odds ratio; \* p<0.05 \*\* p<0.01 \*\*\* p<0.001

## Discussion

The present study demonstrates the pervasiveness of childhood trauma in young people attending mental health services. Approximately 84% of participants reported exposure to at least one form of abuse or neglect during childhood or adolescence, over 67.1% reported experiencing multiple trauma types, and 39% reported trauma in the severe to extreme range. These figures are consistent with trauma reports in a group with early psychosis using a similar trauma measure and a matched control group. In this study 89% of young people with early psychosis reported at least one traumatic experience compared with 37% in the control group and 52% reported at least three traumatic experiences compared with only 7% in the control group (Trauelsen et al., 2015). This suggests that trauma reports, particularly of severe trauma, in the current study would greatly exceed those in the general population. This is supported by evidence that trauma increases the risk for mental disorder in both prospective and meta-analytic studies (Spataro et al., 2004; Widom, 1999; Widom et al., 2007; Cutajar et al., 2010; Varese et al., 2012; Fernandes and Osorio, 2015; Mandelli et al., 2015; Palmier-Claus et al., 2016). The high prevalence of trauma exposure in the young people in the current study shows that trauma is not only common in groups with early psychosis or acute clinical settings (Duhig et al., 2015; Peach et al., 2021; Trauelsen et al., 2015; Allwood et al., 2008; Havens et al., 2012), but extends across help-seeking youth of varying levels of illness severity and mental health concerns.

This high rate of trauma presentation in headspace services highlights the need for a systematic response to trauma in youth mental health services (Bendall et al., 2018). In both the Fifth National Mental Health Plan (COAG Health Council, 2017) and the National Action Plan for the Health of Children in Young People (Australian Government, 2019), the Australian Government has identified that trauma-informed care should be used in all health services to address the needs of those affected by trauma. Trauma-informed care is a service-level approach to providing treatment environments that address the specific needs and sensitivities of trauma-exposed individuals (Substance Abuse and Mental Health Services, 2014). A key limitation of trauma-informed care in youth mental health is the lack of a unified definition and clarity regarding outcomes (Bendall et al., 2021). However, reviewed evidence suggests that there are key clinical interventions that form the basis of any a trauma-informed treatment approach for a wide range of disorders in young people. These include universal screening and assessment, and the consequent provision of psychoeducation for trauma and its effects (Bendall et al., 2021). More work is urgently required to provide an evidence-base for such approaches.

We found a similar pattern of results as Van Nierop and colleagues (2015) in relation to the association between childhood trauma mental health profiles. Childhood trauma was significantly associated with depression, anxiety, mania, and psychosis independently, but was more strongly associated with the co-occurrence of these symptoms than with any one in isolation. These results build on previous research in the following ways. First, they suggest that symptom co-occurrence is already present in trauma-exposed young people with sub-threshold symptoms or emergent mental health disorders. This suggests a proximal effect of childhood trauma on psychopathology, which is consistent with the diverse structural and functional neurobiological changes observed in trauma-exposed youth (Teicher and Samson, 2013). Second, we found trauma-related co-occurrence in simultaneously occurring symptoms rather than lifetime presence. This is important given that concurrent comorbidity has been associated with poor clinical and functional outcomes (Fusar-Poli et al., 2012; Wigman et al., 2012). Finally, the strength of associations between trauma and increasing co-mobidity in the current study were small, as were those found in the non-clinical groups assessed in the study by van Nierop and colleagues (2015). These associations were stronger in adult groups with threschold disorders in the van Nierop and colleagues study, which may suggest that childhood trauma associated symptom admixture becomes more severe over the lifecourse.

The relationship of trauma with more complex symptom presentations in this study underscores the above-described need for a broad approach to treatment rather than intervention for discrete diagnoses. Indeed, recent evidence shows individuals experience multiple disorder types, and that mental health trajectories are better accounted for by age of illness onset, symptom duration, and number of disorder comorbidities than any single diagnosis (Caspi et al., 2020). Further, child maltreatment and interpersonal trauma appear to be important transdiagnostic predictors of heterotypic continuity, accounting for approximately 50% of the risk for transition between mental disorders in a representative US sample (Walsh et al., 2017). We echo calls by van Nierop and colleagues (2015) for further examination of the transdiagnostic efficacy of existing trauma-focused treatments for PTSD. Trauma-focused cognitive behavioral therapy (TF-CBT) is a good candidate in youth mental health, as it was specifically designed for use with children and adolescents, has established efficacy across multiple trauma types and developmental levels, and has been piloted in young people in the Australian headspace setting (de Arellano et al., 2014; Peters et al., 2021; Eastwood et al., 2021). There is also emerging evidence in support of its transdiagnostic efficacy for reducing co-occurring depression and anxiety symptoms (Goldbeck et al., 2016; Lenz and Hollenbaugh, 2017); though these findings are not conclusive (de Arellano et al., 2014). Future research should address these inconsistencies and the dearth of evidence across other symptoms domains, such as early psychosis and early mania, although such work has begun (Tong et al., 2018; Valiente-Gomez et al., 2020). In addition to therapeutic intervention, large scale child abuse and neglect prevention programs and greater support for the child protection system are needed. Such efforts could ultimately be successful in relieving the population burden of mental ill health (Jorm and Mulder, 2018). Existing programs show promise; but quality of evidence and service access for vulnerable groups remain key issues (MacMillan et al., 2009).

The retrospective assessment of childhood trauma in the present study may be a limitation due to the possible influence of recall bias. However, while there is poor to moderate agreement between prospective and retrospective reports of trauma (Baldwin et al., 2019; Reuben et al., 2016; Danese and Widom, 2020) associations with psychopathology have been found irrespective of assessment type (Newbury et al., 2018; Reuben et al., 2016) and data suggests that the retrospective memory of the experience of childhood trauma is a better indicator of psychopathology than documented trauma that is not recalled in adulthood (Danese and Widom, 2020). Further, the effect of retrospectively reported childhood trauma on documented lifetime psychopathology is independent of current psychopathology, suggesting that symptom-related recall bias is not driving differences between retrospective and prospective reports of childhood trauma (Danese and Widom, 2020). Another limitation is the lack of data on post-traumatic stress and negative symptoms of psychosis. Negative symptoms have been associated with child trauma in young people at high risk for psychosis (Yung et al., 2019).

Future studies should collect this information to extend understandings of the interplay between trauma exposure, post-traumatic stress symptoms, and mixed symptomology. Data for this study was collected nearly 10 years ago. However, there have been no major changes to the headspace service model or society in general that would suggest childhood trauma prevalence would have changed in the service setting in those 10 years.

In summary, the present study demonstrates the pervasiveness of childhood trauma and its association with co-occurring depression, anxiety, mania and psychosis symptoms among help-seeking young people. Trauma-exposure appears to be a risk factor for heterogeneous mental health presentations that cut across current diagnostic silos. Mental health practicioners and health services must be cogizant of the high prevalence of trauma and its effects in help-seeking youth and offer appropriate intervention.

## Data Availability

ll data produced in the present work are contained in the manuscript

## Conflict of Interest

The Authors declare that there is no conflict of interest.

## Funding

This work was supported by the National Health and Medical Research Council [grant number 556529].

## Notes

### Competing Interest Statement

The authors have declared no competing interest.

### Funding Statement

This study was funded by the Australian National Health and Medical Research Council [grant number 556529].

### Author Declarations

Ethics committees of the University of Melbourne and the University of Sydney gave ethical approval for this work

